# Single cell sequencing reveals cellular landscape alterations in the airway mucosa of patients with pulmonary long COVID

**DOI:** 10.1101/2024.02.26.24302674

**Authors:** Firoozeh V. Gerayeli, Hye Yun Park, Stephen Milne, Xuan Li, Chen Xi Yang, Josie Tuong, Rachel L Eddy, Elizabeth Guinto, Chung Y Cheung, Julia SW Yang, Cassie Gilchrist, Dina Abbas, Tara Stach, Clarus Leung, Tawimas Shaipanich, Jonathan Leipsic, Graeme Koelwyn, Janice M. Leung, Don D. Sin

## Abstract

To elucidate the important cellular and molecular drivers of pulmonary long COVID, we generated a single-cell transcriptomic map of the airway mucosa using bronchial brushings from patients with long COVID who reported persistent pulmonary symptoms.

Adults with and without long COVID were recruited from the general community in greater Vancouver, Canada. The cohort was divided into those with pulmonary long COVID (PLC), which was defined as persons with new or worsening respiratory symptoms following at least one year from their initial acute SARS-CoV-2 infection (N=9); and control subjects defined as SARS-CoV-2 infected persons whose acute respiratory symptoms had fully resolved or individuals who had not experienced acute COVID-19 (N=9). These participants underwent bronchoscopy from which a single cell suspension was created from bronchial brush samples and then sequenced.

A total of 56,906 cells were recovered for the downstream analysis, with 34,840 cells belonging to the PLC group. A dimensionality reduction plot shows a unique cluster of neutrophils in the PLC group (p<.05). Ingenuity Pathway Analysis revealed that neutrophil degranulation pathway was enriched across epithelial cells. Differential gene expression analysis between the PLC and control groups demonstrated upregulation of mucin genes in secretory cell clusters.

A single-cell transcriptomic landscape of the small airways shows that the PLC airways harbors a dominant neutrophil cluster and an upregulation in the neutrophil-associated activation signature with increased expression of MUC genes in the secretory cells. Together, they suggest that pulmonary symptoms of long COVID may be driven by chronic small airway inflammation.

**Take home message:** Single cell profiling shows the infiltration of neutrophils with upregulation of mucin genes in the airway mucosa of patients with pulmonary long COVID, indicating persistent small airway inflammation in pulmonary long COVID.

## INTRODUCTION

The coronavirus disease 2019 (COVID-19) pandemic has infected more than 771 million people, accounting for more than 6.9 million deaths worldwide [1]. As we transition into the post-pandemic era, a new clinical entity has emerged: post-acute sequelae of severe acute respiratory coronavirus 2 (SARS-CoV-2) infection, commonly known as long COVID [2]. Approximately 10% of all infected adult survivors experience long COVID with almost half reporting persistent symptoms beyond one-year post-infection [3, 4]. However, long COVID is clinically complicated as it is associated with more than 200 different symptoms involving numerous organs in the body [3, 5]. Among the persistent symptoms, respiratory complaints are very common [6–8]. In adult Canadians with long COVID, pulmonary symptoms including dyspnea and cough are reported by 38.5% and 39.3% of all patients, respectively [8]. Despite this, the pathophysiology of pulmonary long COVID remains obscure. To elucidate important cellular and molecular drivers of pulmonary long COVID, we generated a single-cell transcriptomic map of the airway mucosa using bronchial brushings from patients with long COVID and with persistent pulmonary symptoms for more than 1 year following acute SARS-CoV-2 infection.

## METHODS

### Study population

Adults (19 years of age and older) with and without long COVID were recruited from a tertiary care clinic at St. Paul’s Hospital as well as from the general community in greater Vancouver, Canada through advertising. Following informed consent (University of British Columbia/Providence Health Care Research Ethics Board approval H21-02149 and H19-02222), all of the participants answered a series of questionnaires including the St. George’s Respiratory Questionnaire (SGRQ), and underwent pulmonary function test (PFT), low-dose chest computed tomography (CT), hyperpolarized xenon (129Xe) magnetic resonance imaging (MRI) and phlebotomy for complete blood count (CBC). Bronchoscopy was also performed in a subset of these participants who consented to a research bronchoscopy; the details of which have been previously published [9].

*A priori*, the cohort was divided into 3 groups: 1) patients with pulmonary long COVID (PLC), which was defined as persons who remained persistently symptomatic (for >12 weeks post-infection) of new onset or worsening respiratory symptoms including cough or dyspnea, (with a SGRQ total score of >10 units) in the absence of known chronic lung conditions such as bronchiectasis, pulmonary fibrosis or chronic obstructive pulmonary disease (COPD); 2) individuals whose acute respiratory symptoms had fully resolved by 12 weeks of post-infection (i.e. recovered COVID patients); and 3) subjects who had not experienced acute COVID-19 at the time of enrolment. As the demographic and clinical features of groups 2 and 3 were similar, in this study, we merged them as one control group. We excluded any subjects with pre-existing chronic respiratory disorder.

The present cohort study included 24 subjects who underwent bronchoscopy between January 2021 and August 2023. All participants were vaccinated to COVID-19 at the time of enrolment. Based on history, the predominant strains at the time of the patient’s initial SARS-CoV-2 infection were alpha, beta, gamma and delta variants. Bronchoscopies were performed when the subjects were clinically stable for more than 2 months. Among 13 subjects with PLC, 4 subjects were excluded due to poor quality of samples or technical issues in sample processing or library preparation. Among 7 COVID patients without pulmonary symptoms, 2 subjects were also excluded for these reasons. The final number of subjects with PLC in the analysis was 9. The final number of control subjects was also 9 with 5 who had experienced acute COVID-19 but were now free of significant pulmonary symptoms and 4 without a prior history of acute COVID-19 (Figure 1).

**Figure 1:**
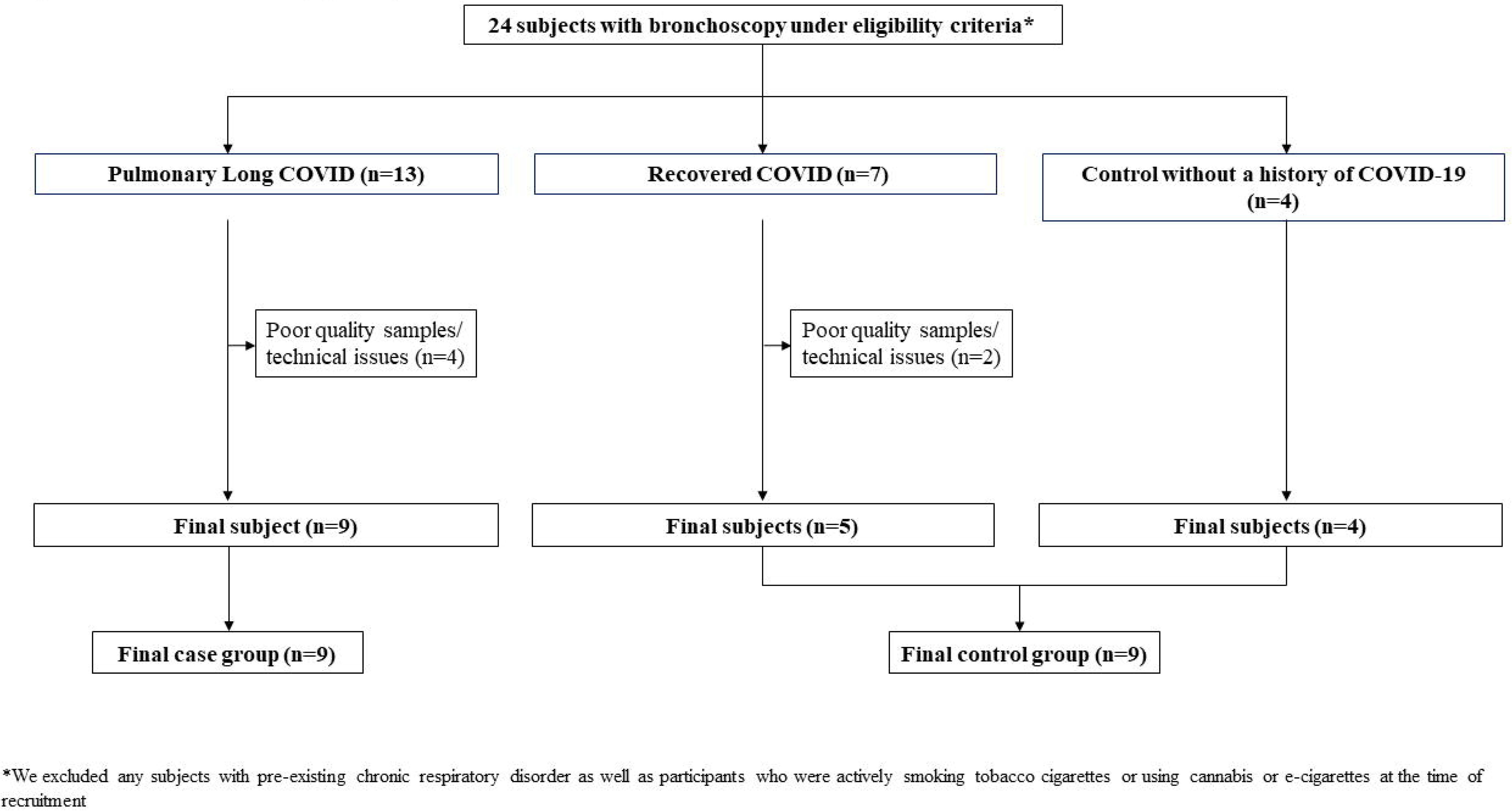
A flow diagram of participants

### Bronchoscopy and single-cell suspension preparation

Under conscious sedation, a fiberoptic bronchoscope (Olympus Corporation, Tokyo, Japan) was passed through the mouth of participants and into the trachea. With the bronchoscope positioned in one of the subsegmental bronchi of the right or left upper lobe, a cytological brush was inserted through a bronchoscope channel into a sixth-to eighth-generation airway from where bronchial brush samples were collected. A cytological brush was then withdrawn from the bronchoscope, and using a pair of stainless steel scissors, the brush was cut into a microcentrifuge tube containing 1000 μl medium and kept on ice until further processing. A single-cell suspension was then created according to our established protocol [10]. In brief, the single-cell suspension resulted from using Accutase dissociation agent (STEMCELL Technologies, Vancouver, BC), and multiple washing steps using Pneumacult-Ex (STEMCELL Technologies, Vancouver, BC) media.

### Single-cell sequencing

The created single-cell suspension was transferred on ice to a sequencing facility where the samples were loaded onto the Chromium Controller using the Chromium Next GEM Single Cell 3′ Kit v3.1 and Chip G (10x Genomics, Inc., CA, USA). The sequenced libraries were prepared in accordance with a previously published Chromium Single Cell 3′ Reagent Kits User Guide [11]. The integrity of the cDNA libraries was examined using a 2100 Bioanalyzer instrument with a High Sensitivity DNA Kit (Agilent Technologies, Inc., CA, USA). Sequencing of the final libraries was performed at a loading concentration of 650 pM with a 2% PhiX spike-in on the NextSeq 2000 (Illumina, Inc., CA, USA) as recommended by 10X Genomics. The final depth of sequenced samples was targeted to reach 60,000 reads/cell. The Fastq files containing the data were generated with Cell Ranger v6.01 (10x Genomics) and the reads were aligned to the human reference genome (hg19).

### Bioinformatics and statistical analysis

Correction for ambient RNA was done using the R package SoupX (version 1.5.2) [12] prior to performing further quality control tasks and downstream analysis. Following ambient RNA correction, using the Pegasus package (version 1.8.1) in Python, we performed additional quality control steps, in which cells with high mitochondrial genes (>20%) or <200 genes were filtered out. To ensure the integrity of the data, we also filtered out cells that expressed > 8000 genes, as very high gene counts could be the result of contamination or a technical artifact. As the last step in data quality control, we removed cells with >1% hemoglobin gene expression along with those that expressed MALAT1, mitochondrial, or ribosomal genes. This method was applied to reduce the impact of the aforementioned genes on downstream analysis and to improve the accuracy of gene expression profiles. The data were then processed for normalization, logarithmic transformation, and batch effect correction using the Scanorama algorithm (version 1.7.3). During this process, doublets were removed through the Pegasus package. Cell type annotation was performed predominantly using the Pegasus human lung and human immune legacy markers, which was complemented by other resources including; the Protein Atlas, PanglaoDB [13, 14] and the methods by Hewitt et.al and Zaragosi et.al (Supplemental Table S1) [15, 16]. Wilcoxon rank-sum tests were used to determine if there were significant cell proportion differences between the PLC group and controls. We also performed differential gene expression between the two groups, and significantly expressed genes were identified at a false discovery rate (FDR) of < 0.05. Cell trajectory and pseudotime analysis were carried out using Velocyto 0.17.17, and ScVelo 0.2.5 packages in Python. All analyses were performed in R (4.3.1) and Python (3.9.12).

Ingenuity Pathway Analysis (IPA) was also conducted on differentially expressed genes discovered based on a False Discovery Rate (FDR) of 1.0. The gene lists from each cell cluster were then combined into an Excel file and uploaded onto QIAGEN IPA (version 01-22-01). Using the gene names and Ensemble IDs, the genes are then mapped onto the QIAGEN IPA server and an expression core analysis was performed using an Experimental Log Fold Change. User Dataset was the population of genes based on which the p-values were recalculated with both direct and indirect relationships. The absolute value of 0.5 and 0.05 was set as the Experimental Log Ratio cut-off and the Experimental False Discovery Rate (q-value), respectively. QIAGEN IPA uses the input parameters (Log ratio cut-off and q-value) to calculate a z-score, which was then used to infer the pathway’s activity. Orange colour indicates pathways that have positive z-scores and are predicted to be activated. Blue colour indicates pathways that have negative z-scores and are predicted to be inhibited. White colour indicates pathways that have a z-score equal to 0 and gray colour indicates pathways that have activity, but their activity cannot be predicted. We selected the top 30 upregulated pathways to examine the pathways activated in PLC compared to controls.

## RESULTS

### Study subjects

There were no significant differences in age, sex, smoking status, or pulmonary function test measures between subjects with PLC (n=9) and controls (n=9). However, the median (IQR) body mass index (BMI) was significantly higher in subjects with PLC than in control subjects [28 kg/m2(27 – 31) vs. 23 (22 - 26.03); *p*=0.03]. Similarly, the median (IQR) SGRQ total score and all three domains of symptoms, activity and impact were also significantly higher in subjects with PLC than in control subjects [45.0 (33.1 -69.1) vs. 2.1 (1.0-5.7); *P* < 0.001 for SGRQ total score, 42.7 (40.8-44.1) vs. 5.7 (0-8.8); *P* < 0.001 for Symptoms, 60.4 (48.3 -86.5) vs. 0 (0-12.2); *P* < 0.001 for Activity domain and 32.49 (20.97-64.55) vs. 0 (0-0); *P* < 0.001 for Impact domain] (Table 1) There were no significant differences in blood leukocytes or their differentials including neutrophils, lymphocytes, monocytes, eosinophils and basophils between the two groups. In bronchoalveolar lavage fluid, the cell counts for alveolar macrophages, lymphocytes, neutrophils and eosinophils were similar between the two groups and there was no evidence of a microbiologic infection (Table 1). The median (IQR) follow-up days from COVID-19 to bronchoscopy was 711 (444 - 832) and 556 (374 - 703) for PLC group and control group (recovered COVID patients), respectively. On a low-dose chest CT scan, there were no abnormal findings in the control group and there were minimal abnormal findings in the PLC group, which included mosaic attenuation (n=1), ground glass opacity (n=1) and reticulation (n=3) without emphysema or honeycombing (Table 2).

**Table 1.** Baseline characteristics of study participants.

|  | Pulmonary Long COVID (n=9) | Control (n=9) | P value |
| --- | --- | --- | --- |
| Age, years | 54 (44 - 60) | 35 (24 - 62) | 0.62 |
| Male, n (%) | 5 (55.6%) | 4 (44.4) | 1.00 |
| BMI, kg/m<sup>2</sup> | 28 (27 - 31) | 23 (22 - 26) | 0.03 |
| Tobacco Smokers |  |  | 1.00 |
| former | 1 (11.1) | 1 (11.1) |  |
| Never | 8 (88.9) | 8 (88.9) |  |
| Cannabis Smokers |  |  | 0.47 |
| Current | 0 | 2 (22.2) |  |
| Never | 9 (100) | 7 (77.8) |  |
| E-cigarette Users |  |  | 1.00 |
| Current | 0 | 1 (11.1) |  |
| Never | 9 (100) | 8 (88.9) |  |
| SGRQ, total | 45.0 (33.1 - 69.1) | 2.1 (1.0 - 5.7) | <0.001 |
| Symptoms | 42.7 (40.8 - 44.1) | 5.7 (0 - 8.8) | <0.001 |
| Activity | 60.4 (48.3 - 86.5) | 0 (0 - 12.2) | <0.001 |
| Impact | 32.49 (20.97 - 63.55) | 0 (0 - 0) | <0.001 |
| Pulmonary Function Test |  |  |  |
| FVC, L | 3.99 (2.96 - 5.42) | 3.97 (3.84 - 4.77) | 1.00 |
| FVC, % of predicted | 108.6 (101.61 - 114.62) | 117.8 (106.6 - 126.5) | 0.19 |
| FEV<sub>1</sub>, L | 2.97 (2.43 - 4.23) | 3.42 (3.01 - 4.08) | 0.48 |
| FEV<sub>1</sub>, % of predicted | 110.7 (94.7 - 117.1) | 117.7 (106.3 - 125.0) | 0.11 |
| FEV<sub>1</sub>/FVC (%) | 80.6 (76.8 - 84.3) | 81.0 (79.8 - 86.9) | 0.66 |
| DLco, % of predicted | 96.9 (83.8 - 102.5) | 108.1 (100.7 - 112.2) | 0.13 |
| Blood cell counts |  |  |  |
| White blood cells (x 10<sup>9</sup>/L) | 5.28 (4.74 - 6.48) | 4.63 (4.49 - 5.11) | 0.31 |
| Neutrophils (%) | 56.4 (52.05 - 58.45) | 55.7 (53.9 - 61.7) | 1.00 |
| Lymphocytes (%) | 32.5 (30.35 - 38.5) | 32.3 (28.4 - 34.9) | 0.60 |
| Monocytes (%) | 5.1 (3.95 - 6.4) | 4.3 (4.1 - 4.9) | 0.60 |
| Eosinophils (%) | 2.2 (1.6 - 3.05) | 2.0 (1.9 - 4.2) | 0.50 |
| Basophils (%) | 0.6 (0.45 - 0.9) | 0.5 (0.4 - 0.7) | 0.60 |
| Bronchoalveolar lavage |  |  |  |
| Alveolar macrophage, (%) | 79 (76 - 95) | 86 (77 - 93) | 0.79 |
| Lymphocytes (%) | 18 (4 - 20) | 8 (4 - 16) | 0.59 |
| Neutrophils (%) | 1 (0 - 3) | 2 (1 - 5) | 0.37 |
| Eosinophils (%) | 0 (0 - 0) | 0 (0 - 0) | 0.37 |
| Days from COVID-19 to bronchoscopy | 711 (444 - 832) | 556 (374 - 703)* | 0.35 |

Definition of abbreviations: BMI = body mass index; DLco =diffusion capacity for carbon monoxide; FVC = forced vital capacity; FEV<sub>1</sub> = forced expiratory volume in 1 s; MPXI = myeloperoxidase index; SGRQ = St. George's Respiratory Questionnaire.
\* The time from infection to bronchoscopy for individuals in the control group that are post COVID but do not report any persistent pulmonary symptoms (n=4)
Data are expressed as median (interquartile range) or n (%).

**Table 2.**
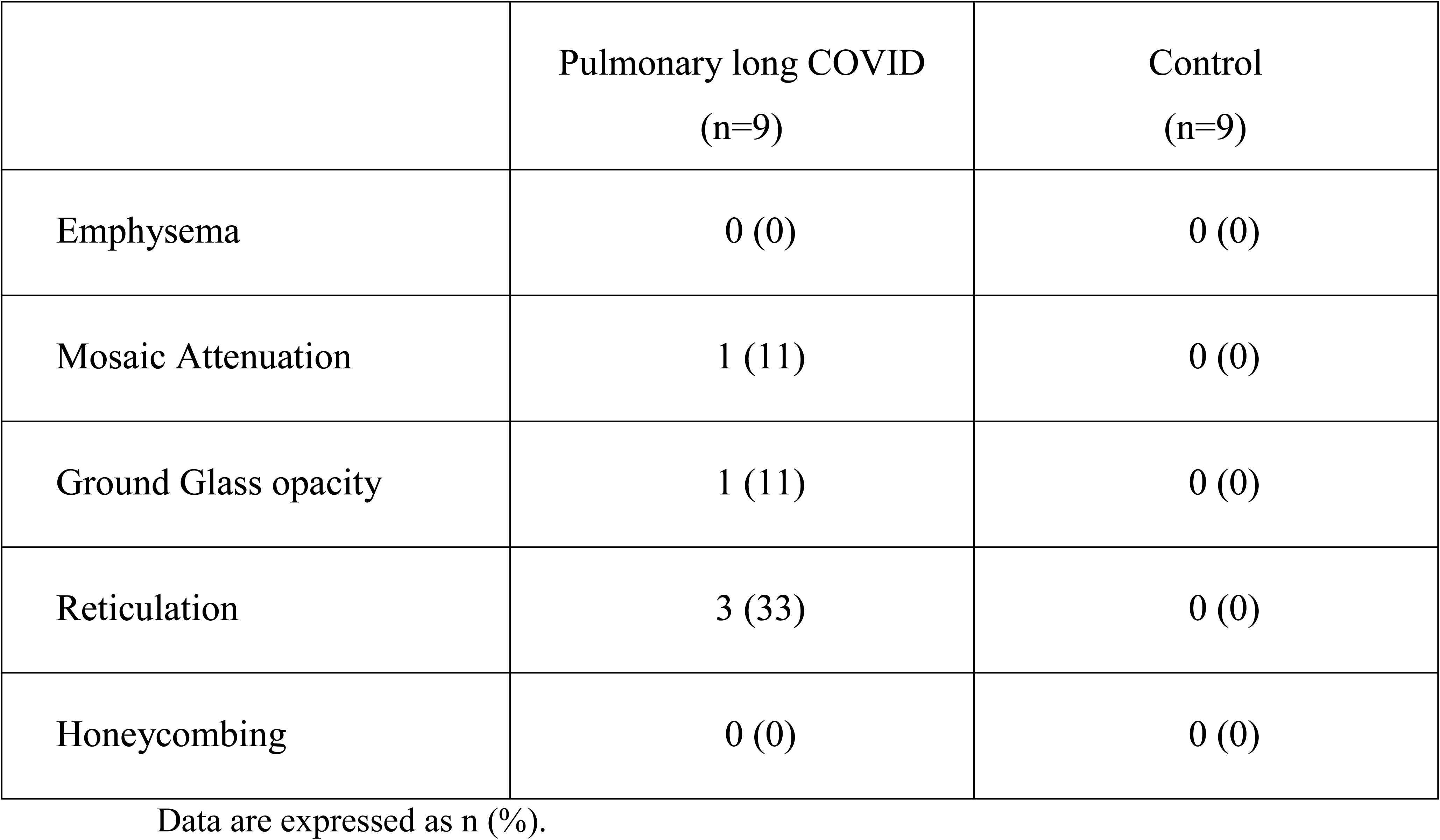
Chest computed tomography findings.

### Profiling the mucosal landscape using single-cell RNA sequencing

A total of 56,906 cells were recovered for the downstream analysis, with 34,840 cells belonging to the PLC group. A dimensionality reduction plot using Uniform Manifold Approximation (UMAP) demonstrated several clusters that were identified by using legacy markers and manual curation (Figure 2). Using this approach, we identified a cluster of cells that were predominantly derived from bronchial brush samples of the PLC patients (Figure 2A), and noted 16 clusters that were of epithelial origin and 15 clusters that were immune cells (Figure 2B). A composition plot (Figure 3) shows the distribution of the aforementioned clusters. A Wilcoxon test revealed that, when comparing PLC and control group across all clusters, the neutrophil cluster exhibited statistical significance (p<.05).

**Figure 2:**
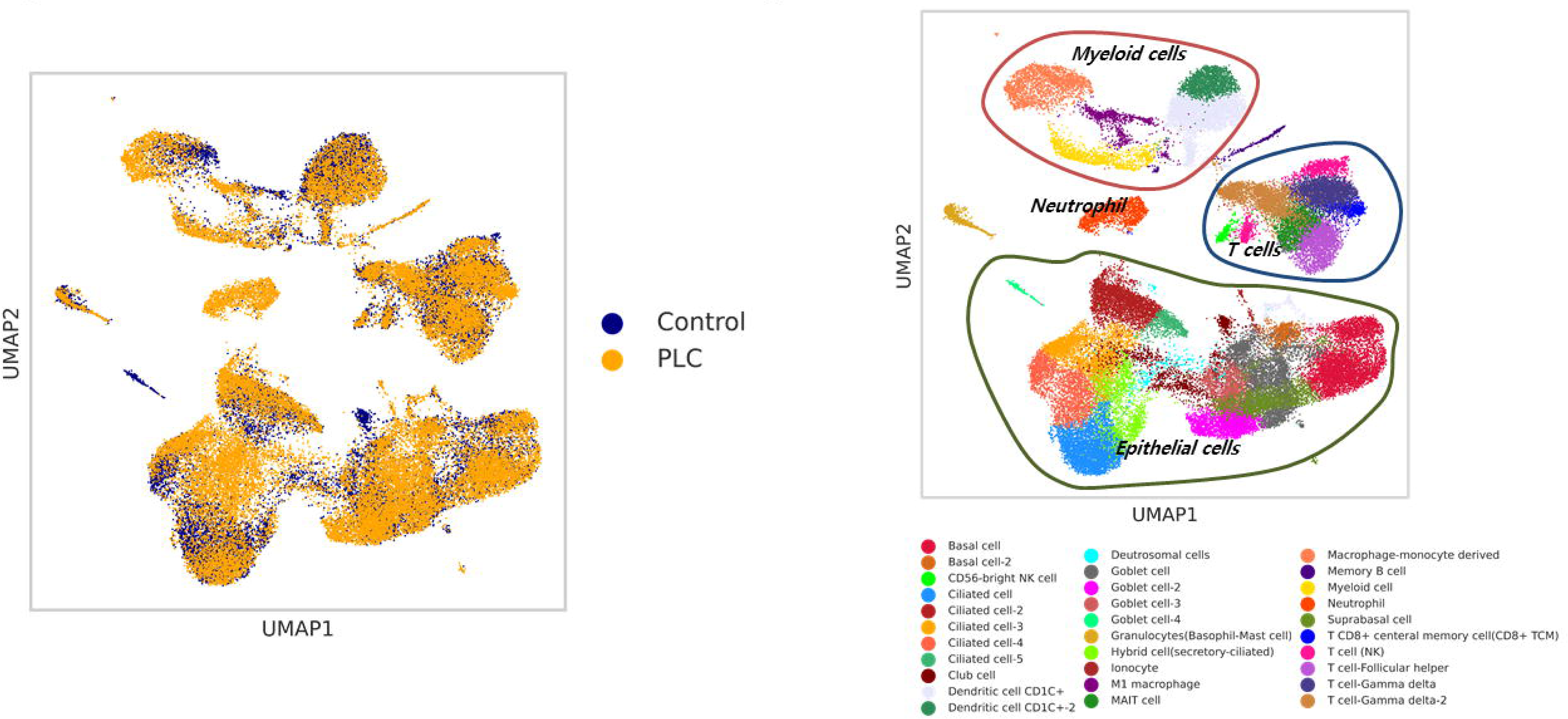
Uniform Manifold Approximation (UMAP) of 56,906 of profiled cells from both PLC and control group, which were annotated using legacy markers. A: A dimensionality reduction plot of PLC versus control reveals distribution of clusters across both categories. It prominently highlights a specific cluster derived from the PLC group. B: Various clusters of cells originating from both epithelial and immune cell lineages were observed. As anticipated, the diversity among these identified cell clusters serves as a representation of the cellular heterogeneity within the airway mucosa. Additionally, distinct neutrophil cluster was highlighted.

**Figure 3:**
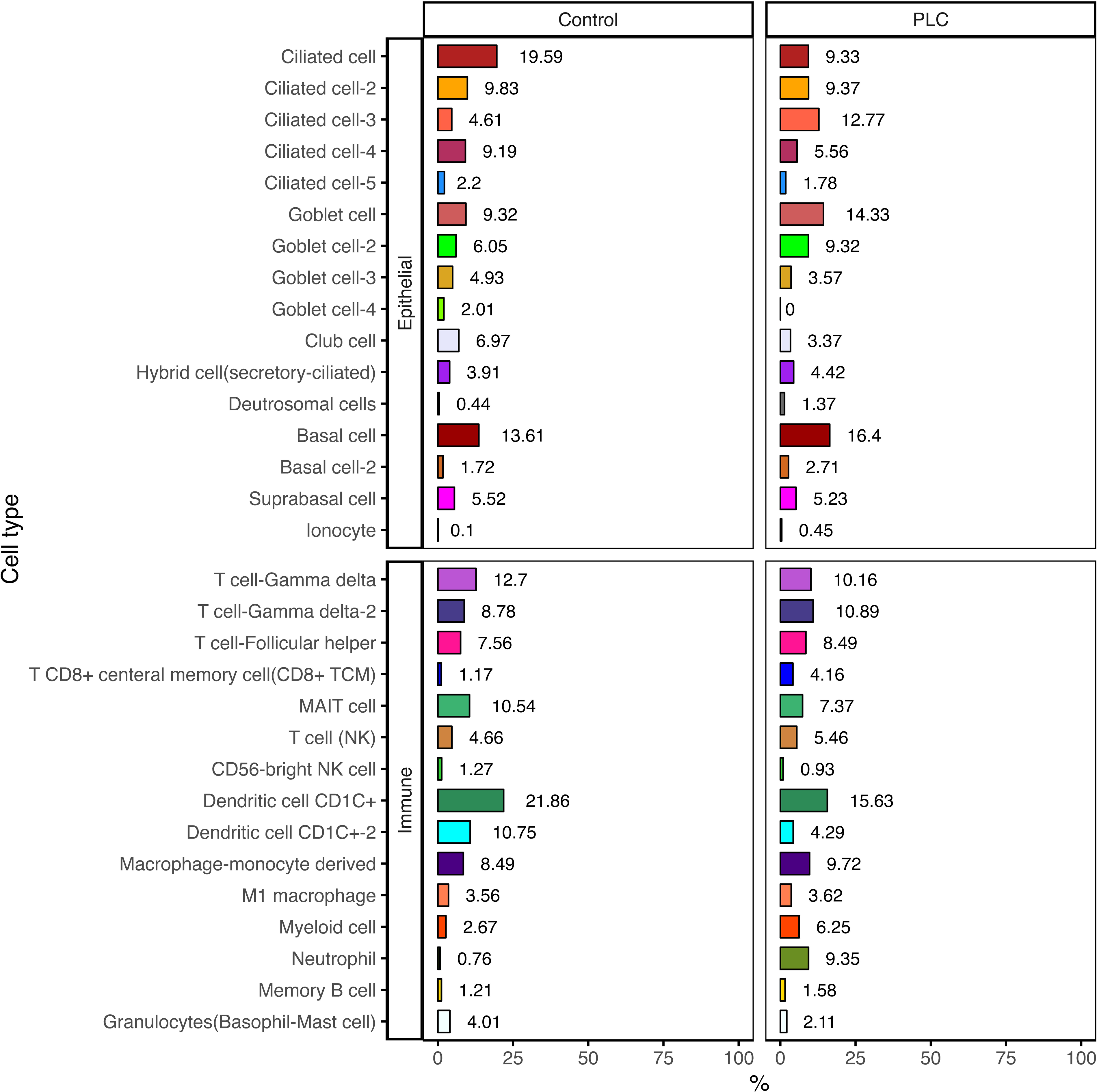
A composition plot showing different cell types identified in the sample.

### Trajectory analysis

The RNA velocity dynamical model in conjunction with a pseudotime analysis revealed that a majority of cells in our dataset were relatively transcriptomically stable and thus likely in a quiescent state. This was confirmed by evaluating the Antigen Kiel 67 (MKI67) expression levels across all cell types (Supplemental Figure S1A-B). The calculated coherence of the vector field provided us confidence of accuracy for the calculated RNA velocity (Supplemental Figure S1C)

### Differential gene expression analysis of epithelial and immune cells

Compared to control subjects, transcriptional profiling of MUC genes revealed upregulation of MUC1, MCU5AC and MUC5B across PLC epithelial cells with secretory characteristics (Figure 4A-B) compared to controls.

**Figure 4:**
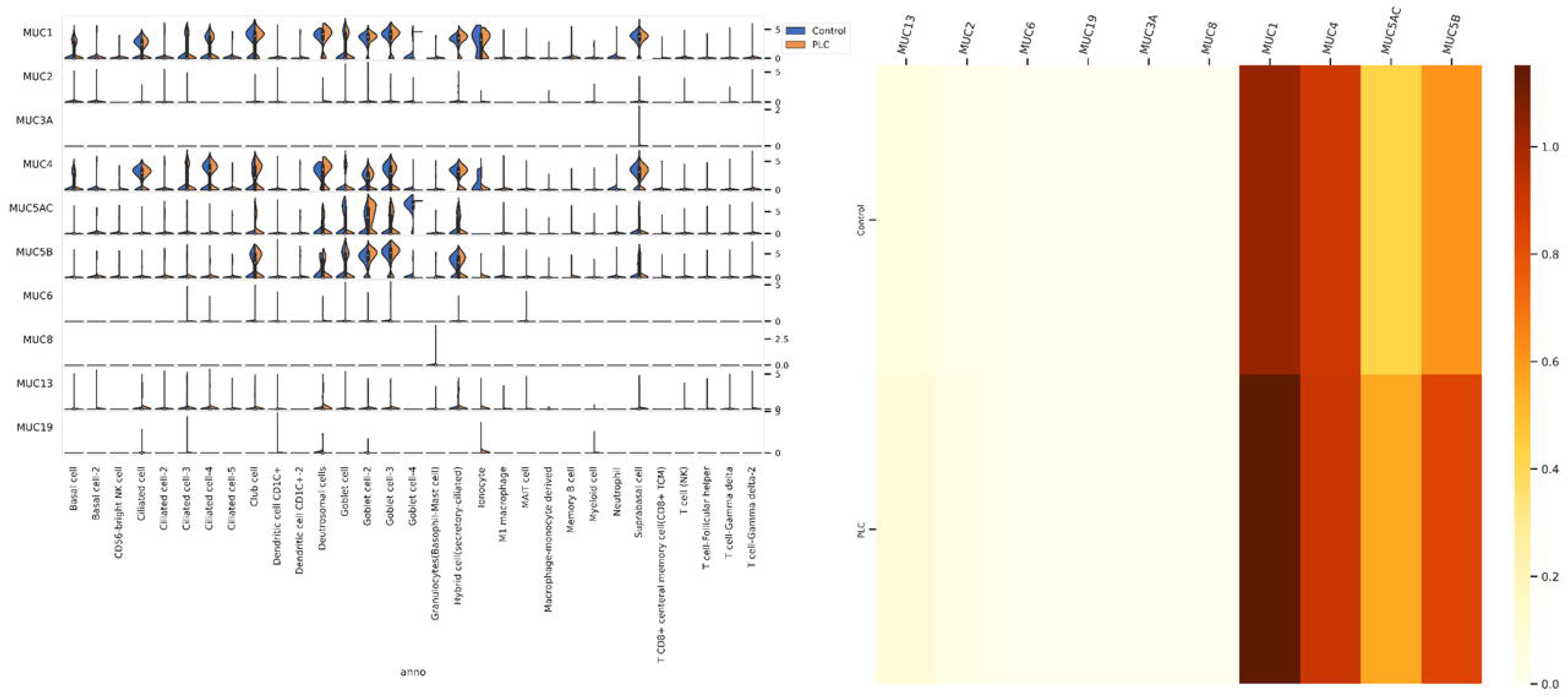
MUC genes expression across identified cell types: A: A violin plot showing average MUC gene expression across various PLC and control clusters. B: A heatmap indicating differences in the average expression of various MUC genes in PLC vs controls.

Differentially expressed genes between PLC and control participants were subjected to comparative canonical pathway analysis in QIAGEN IPA. Here, we focused on differentially expressed genes in secretory cells from PLC group (i.e. goblet cell, goblet cell-2 and club cells) that had higher expression of MUC genes. We also explored canonical pathway analysis in IPA of basal cells to understand if neutrophil infiltration perturbs these progenitor cell homeostasis. Both secretory and basal cells consistently showed enrichment of pathways associated with neutrophil degranulation. Additionally, in secretory cells, interleukin (IL)-13, and T cell receptor signaling was enriched based on the IPA analysis (Figure 5A-C), whereas basal cells demonstrated pathway upregulation associated with neutrophil extracellular trap signaling (Figure 5D). We refrained from differential gene analysis on the neutrophil cluster between PLC and controls, since a vast majority of the cell constructing this cluster originated from the PLC group, hence any differential analysis would be skewed. The IPA also showed potential chemicals and compounds that may reverse the transcriptomic signature in various cells (Supplemental Figure S2). For example, in club cells, budesonide is predicted to revert the signature of PLC airways to that resembling the control airway.

**Figure 5:**
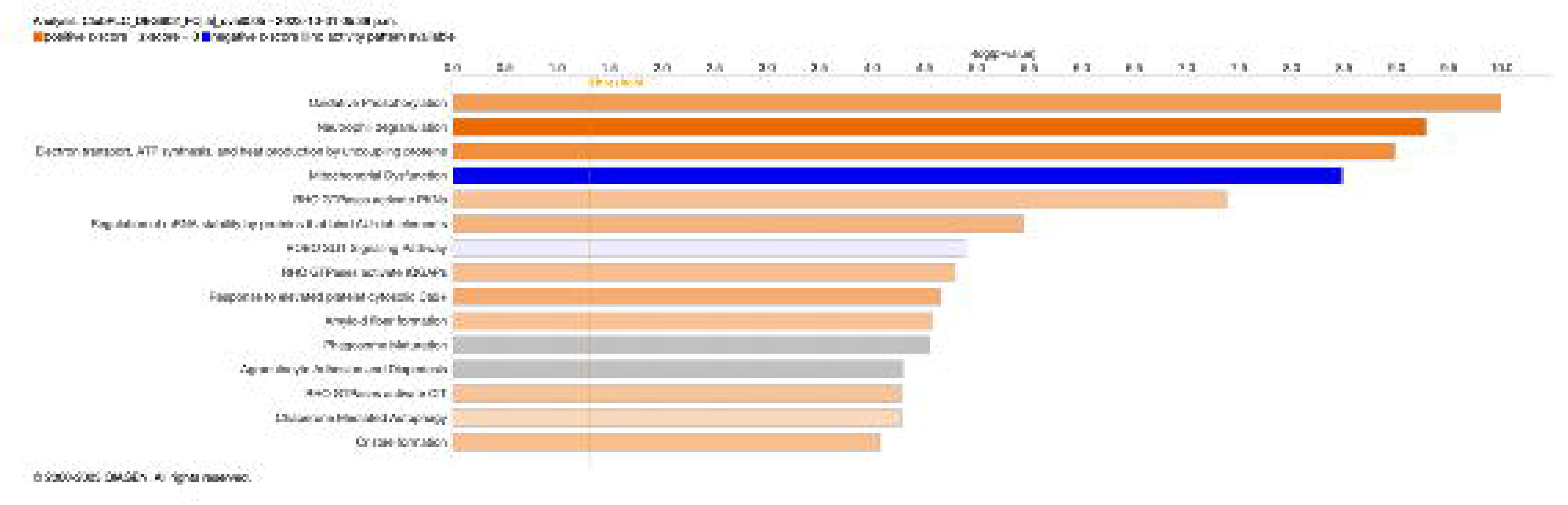
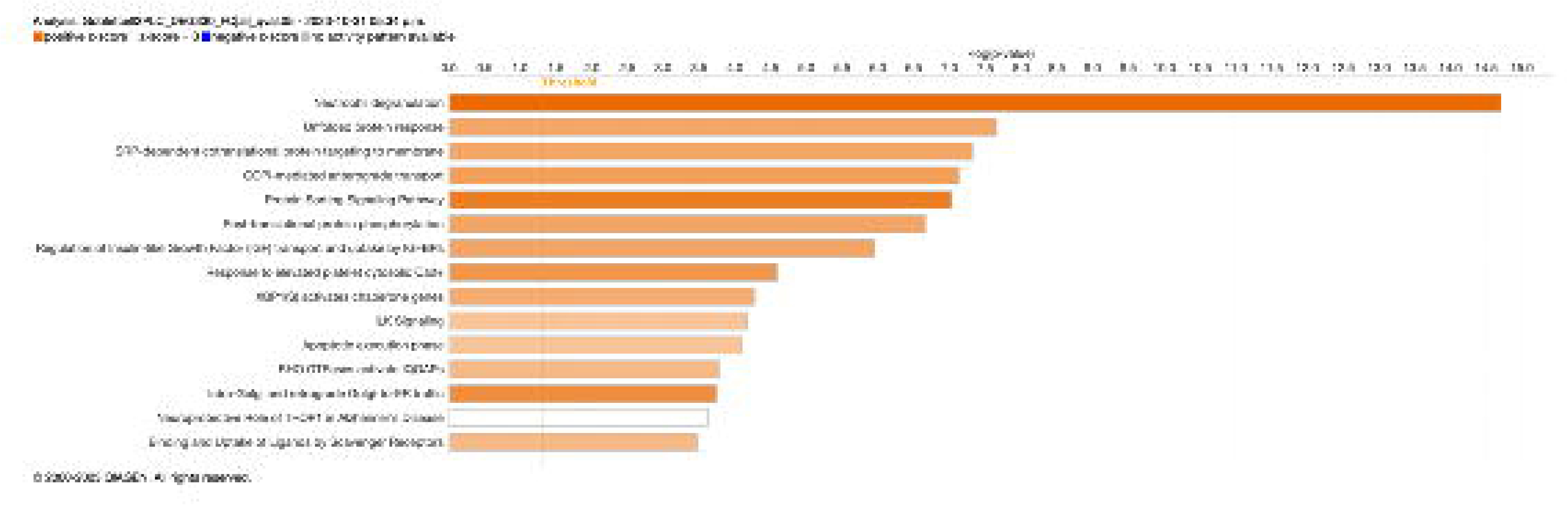
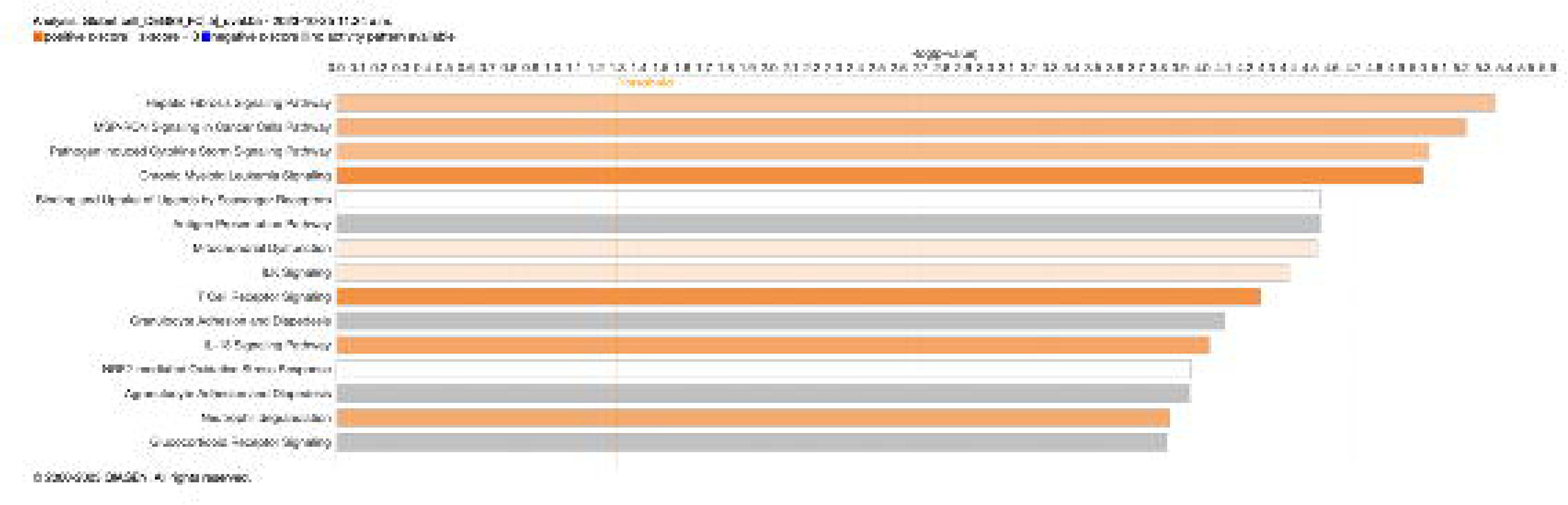
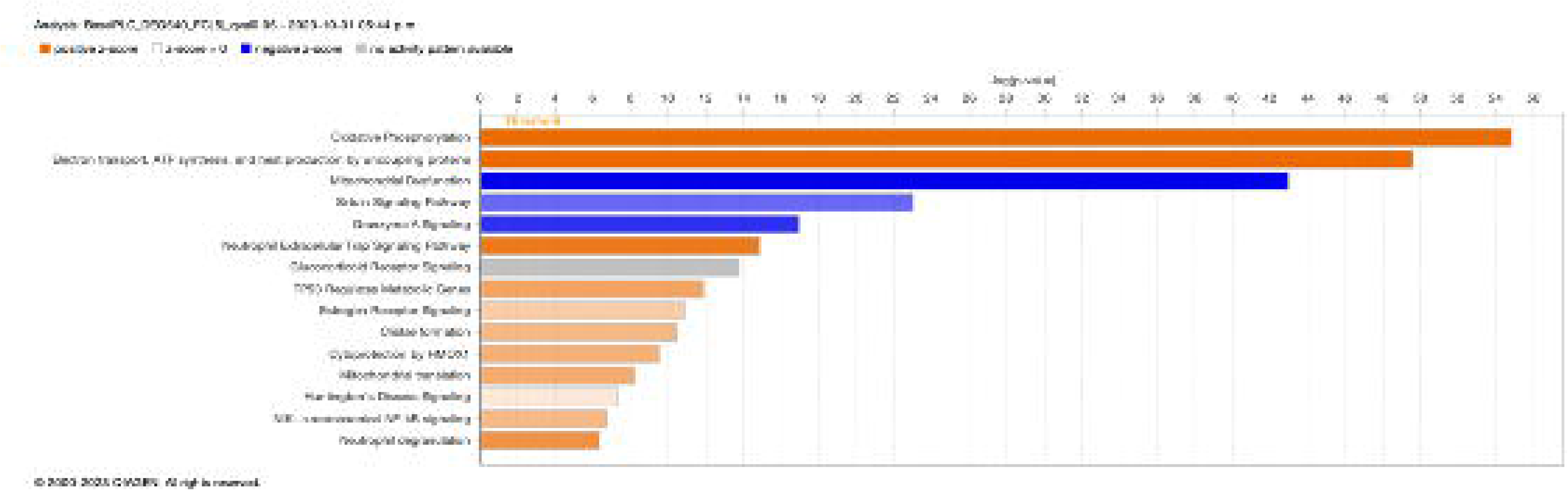
IPA analysis shows enrichment of various pathway across A: Club cell B: Goblet cell C: Goblet cell-2 D: Basal cell

Among the 5 most common cell types (ciliated, secretory, dendritic, T and myeloid lineage cells), we found 95 common genes across all five clusters that were upregulated in PLC samples (Figure 6). These genes were involved in various inflammatory pathways, such as response to viruses, response to interferon-alpha/beta and gamma and neutrophil mediated immunity.

**Figure 6:** A: Venn Diagram of the most abundant cell types where 93 genes were identified to be overlapping between them. B: A table showing GO pathways associated with these genes.

## DISCUSSION

Here, we profiled the transcriptomic landscape of epithelial and airway immune cells in the small airways of patients with PLC at a single cell resolution. The most striking feature was an increase in the number of neutrophils in the airway mucosa as well an upregulation in the neutrophil-associated activation signatures across clusters in the PLC airways. We also observed an increase in MUC gene expression in the secretory cells of the PLC airways.

Our findings are in line with the important role neutrophils play in the pathogenesis of severe COVID-19. During an acute infection, there is a significant increase in circulating neutrophils in peripheral blood of patients with severe COVID-19 infections [17, 18] and several studies using scRNA-seq have reported an increase in dysfunctional circulating neutrophils in the blood of those with severe disease [19, 20], which together may contribute to cytokine storm caused by uncontrolled innate immune system activation during active disease [21, 22].

The pathogenesis of long COVID-19, however, is largely unknown. A previous study indicated that individuals with interstitial lung changes at 3 to 6 months post-infection demonstrated upregulation in neutrophil-associated immune signatures including increased chemokines, proteases, and markers of neutrophil extracellular traps in circulation compared to those who experienced complete radiographic resolution at follow-up [23]. A recent study showed that long COVID patients display up-regulation of certain inflammatory plasma proteins. Interestingly, the most enriched pathways (for these proteins) were those related to neutrophil degranulation [7]. We extend these data by showing that neutrophils and their associated signatures are increased in the small airway mucosa of patients with PLC, even among patients with no or minimal changes on chest imaging (by CT scan) or PFTs. Additionally, IPA and other bioinformatics analysis showed enrichment of neutrophilic degranulation pathways in both secretory and basal cells, while neutrophil extracellular trap signaling pathway was enriched in basal cells. These data suggest that even months and years following acute COVID, there may be “neutrophilic” inflammation in the small airways of long COVID patients, who have persistent pulmonary symptoms.

The upstream drivers of the inflammatory changes in the airway mucosa of PLC patients are obscure and largely speculative. Given that lung epithelial cells are the prime targets of entry and propagation of SARS-CoV-2, the enrichment of adaptive immune response signaling (IL-13, and TCR signaling) in ciliated cells raises the possibility of ongoing stimulation potentially by putative viral reservoirs [24, 25]. However, we did not find any evidence in the airway tissue for viral particles or genome (data not shown). Another possibility was raised by a previous study, which showed patients with long COVID display an exhausted SARS-CoV-2-specific T-cell response (CD8+ CD28−) even in mild cases of COVID-19 [26]. Exhaustion of adaptive immunity may skew the immune response towards a dysregulated innate immunity, leading to localized inflammation. Notably, we saw an enrichment of T cell receptor signaling and IL-13 signaling pathway in goblet cells as well as neutrophilic degranulation pathway. IL-13 induces goblet cell hyperplasia and mucus production in airway epithelial cells [27, 28]. In line with these observations, we saw increased expression of MUC5AC and MUC5B genes, representing two major secreted airway mucins, the airways of PLC compared with control subjects [29]. We also cannot discount the possibility of auto-antibodies or changes in the local microbiome as potential upstream drivers of the inflammatory changes in the PLC small airways.

Dissimilar to prior studies, which used circulating tissues for scRNAseq, we used airway tissue samples obtained during research bronchoscopy. Moreover, we focused on patients with long COVID who had persistent pulmonary symptoms for longer than one year post-acute infection, which enabled us to relate the scRNAseq data in the small airways of these patients with their symptoms. It is notable that these patients had normal lung function measurements and normal or near normal CT imaging.

The present study has several limitations. First, this is a cross-sectional study, thus longitudinal changes of involved cells and biological pathway for PLC could not be investigated. Secondly, due to the nature of single-cell transcriptomic analysis, the expression level of protein related to function could not be assessed. Thirdly, technical limitations of scRNA sequencing such as dropout events of fragile cells and relatively low sequencing depth, leading to the loss of rare cell types or those with low RNA content could also have skewed our results, though any confounding or biases from these technical issues should have been non-differential and affected both the PLC and control groups, resulting in a reduction in the signal. Finally, our primary recruitment relied on voluntary participation from the local community, thus, the results of our study may not be generalizable to other different settings.

In conclusion, a single-cell transcriptomic landscape showed an increase in the number of neutrophils and an upregulation in the neutrophil-associated activation signature in the PLC airways of patients who were more than one year post-acute infection. An increase in MUC gene expression in epithelial cells of PLC airways was also observed. Together, these changes may explain the persistent pulmonary symptoms of cough, sputum production and exertional dyspnea, though additional studies will be required to fully validate this notion. Notwithstanding, these chronic inflammatory changes in the small airways provide new biologic insight on the pathogenesis of PLC and raise novel (potential) therapeutic investigations for these patients.

## Data Availability

All data produced in the present study are available upon reasonable request to the authors

## Supplemental Figures

**Supplemental Figure S1A-C:** RNA velocity and pseudotime analysis show majority of cells being in quiescent state.

**Supplemental Figure S2:** Potential therapeutic targets recognized by IPA pathway analysis of secretory cell clusters.

